# Precise blood proteome profiling in an undiagnosed population with COVID-19

**DOI:** 10.1101/2021.11.15.21266315

**Authors:** Claudia Fredolini, Tea Dodig-Crnković, Annika Bendes, Leo Dahl, Matilda Dale, Cecilia Mattsson, Cecilia E Thomas, Olof Beck, Niclas Roxhed, Jochen M Schwenk

**Affiliations:** Science for Life Laboratory, School of Engineering Sciences in Chemistry, Biotechnology and Health, KTH Royal Institute of Technology, 171 65 Solna, Sweden; Micro and Nanosystems, School of Electrical Engineering and Computer Science, KTH Royal Institute of Technology Stockholm, 100 44 Stockholm, Sweden; Department of Clinical Neuroscience, Karolinska Institutet, 171 77 Stockholm, Sweden; MedTechLabs, BioClinicum, Karolinska University Hospital, 171 64 Solna, Sweden

## Abstract

Self-sampled blood provided valuable information about the COVID-19 seroprevalence in the general population. To enable an even deeper understanding of pathophysiological processes following SARS-CoV-2 infections, 276 circulating proteins were quantified by proximity extension assays in dried blood spots (DBS). Samples from undiagnosed individuals collected during the first wave of the pandemic were selected based on their serological immune response and matched on self-reported symptoms. We stratified these as seropositive (IgM^+^IgG^+^; *N* = 41) or seronegative (IgM^-^IgG^-^; *N* = 37), and to represent the acute (IgM^+^IgG^-^; *N* = 26) and convalescent phases (IgM^-^IgG^+^; *N* = 40). This revealed proteins from a variety of clinical processes including inflammation and immune response (MBL2, MMP3, IL2RA, FCGR2A, CCL5), haemostasis (GP1BA, VWF), stress response (ANG), virus entry (SDC4) or nerve regeneration (CHL1). The presented approach complements clinical surveys and enables a deep molecular and population-wide analysis of COVID-19 from blood specimens collected outside a hospital setting.

## Introduction

After almost two years into the COVID-19 pandemic and more than two hundred million infected people, there is still much to learn about a coronavirus infection resulting in a wide range of clinical manifestations. Initially considered a disease of the respiratory system, the list of symptoms observed in COVID-19 patients has grown over the time, revealing damage to all major physiological functions from the cardiovascular, digestive and nervous systems (*1*). A key factor in COVID-19 pathogenesis is the hyper activation of the innate immune response with consequent cytokine storm (*2, 3*).

In the first months of the pandemic emergencies, focus was to better understand and treat severe COVID-19, to identify effective therapies, and to reduce the mortality. Indeed, great progresses have been made in understanding the molecular mechanism behind severe disease in the critically ill and in the development of vaccine, however still little is known about the long-term effects in those with mild or asymptomatic forms of COVID-19. Symptoms such as severe fatigue, memory lapses, and cardiovascular problems have been found for patients that were mildly affected, especially when symptoms last over a longer period of time and hindering a full recovery (*4*). It has also been shown that even asymptomatic and mild symptomatic infection may be associated with subclinical lung abnormalities, as detected by computed tomography (5, 6). Population-based studies to better understand the heterogeneous phenotypes as well as genetic and environmental factors associated with disease risk and mortality, long-term effects on individual’s well-being, and to identify therapies that address the molecular diversity are consequently urgently needed (7, 8). Large-scale population studies have been initiated, but these may be hampered by an inclusion bias. The practical challenges lie on the inclusion of a general population of undiagnosed or non-hospitalized individuals (affected by mild or no symptoms).

One possibility to engage a wider range of phenotypes is to use home-sampled dried blood spots (DBS). This strategy can facilitate a sample collection across hard-to-reach population groups and reduce the risk to the bias in the study design (*9*). DBS sample collection is in use since the 60’s and has a large usage in clinical chemistry applications particularly for newborn screening. Advantages of DBS collection in respect to traditional blood sampling include (i) not requiring direct contact and expertise of healthcare personnel, (ii) avoid travelling and visiting a healthcare centre; (iii) representing a convenient format for storage and transportation, and (iv) reduced cost both from a societal and healthcare perspective(*10*). However, the haematocrit-bias and imprecision of the collected blood volumes has been issues that hindered an even wider use of DBS in medical practice. These two drawbacks have, however, been recently overcome by new microfluidic-based DBS devices that enable a precise volume-controlled blood collection (*11-15*).

During the first wave of the pandemic, we used a volumetric DBS home-sampling device to collect 878 samples from random households in Stockholm, Sweden. We determined the presence of IgG and IgM antibodies against several proteins from the SARS-CoV-2 virus in a multi-analyte assay (*16*). An accumulated seroprevalence of 10% and available questionnaire data prompted the question to further define molecular features of seropositive status. Therefore, here we used DBS samples collected in the Stockholm population to perform additional analyses to deepen our understanding of the molecular effects of the infection. We profiled 276 circulating proteins involved in cardiovascular and metabolism functions in eluates prepared previously for the serological survey (*16*). The aim of this study was to determine a comprehensive picture of both the multiple biological processes associated with SARS-CoV-2 infection, as well as the diverse facets of the human immune response in a random population.

## Results

Using a volumetric microfluidic-based DBS device that collects exactly 10 μl of whole blood, a protocol was tailored for the analysis of 276 proteins by proximity extension assays (PEA) (**Fig. 1**). After benchmarking the procedure in a pilot study against paired EDTA plasma samples, DBS collected in the Stockholm population during spring of 2020 were analyzed for proteins associated with SARS-CoV-2 seropositivity. The study revealed proteins relevant to COVID-19 pathogenesis and immune response.

**Fig. 1.**
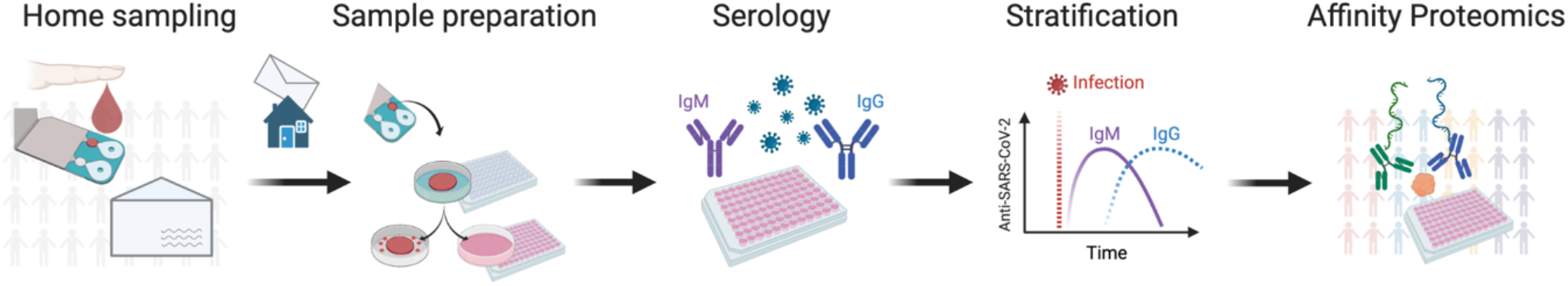
Proteomic profiling of population-based DBS samples. Home-sampling devices were distributed by postal mail to random individuals in metropolitan Stockholm during spring 2020. Dried blood spots (DBS) were collected by finger pricking and mailed back to our laboratory for analysis. We eluted proteins from the DBS discs to first determine antibodies against the SARS-CoV-2. Stratifying by serostatus, we applied proximity extension assays (PEA) to measure the levels of 276 proteins in relation to the immune response.

### Comparison of EDTA plasma and DBS eluates supports the suitability of proteome analysis of DBS samples

To assess the suitability of the DBS preparation for proteomics analyses, protein profiles of 92 circulating proteins related to cardiovascular diseases were investigated **(Figs. 2A** and **B**). The response levels and correlation between proteins detected were compared between EDTA plasma collected by venous blood draw and corresponding DBS samples collected at the same visit by finger-pricking from 12 donors (**Data S1)**. In total, 91 out of 92 proteins were detected in > 90% of the sample types, respectively. The investigated proteins could therefore be measured in DBS and paired plasma samples. When searching for proteins with different abundance levels (FDR *P* < 0.01) as shown in **Fig. 2A**, we found proteins more abundant in DBS with platelet glycoprotein VI (GP6), bleomycin hydrolase (BLMH), azurocidin 1 (AZU1) and caspase 3 (CASP3) being top ranked. Likewise, we found proteins more abundant in plasma with COL1A1 top ranked (**Data S1**). We also correlated the protein profiles to compare the ranking of the paired samples 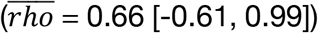, see **Fig. 2B**. In general, 62% (57/92) of the proteins presented with an elevated inter-sample correlation (*rho* > 0.7). Protein profiles of CASP3, PRTN3, JAMA and SELP were most discordant (*rho* < 0), while NPPB, IGFBP1, EPCAM, LDLR, CD163, CPB1, and SELE were highly concordant (*rho* > 0.95) between plasma and DBS. Profiles of proteins elevated in plasma agreed with DBS profiles 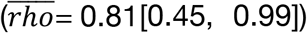 However, profiles of proteins elevated in DBS did agree less with the respective plasma profiles 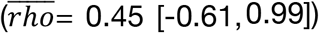, see **Fig. S1** and **Data S1**.

**Fig. 2.**
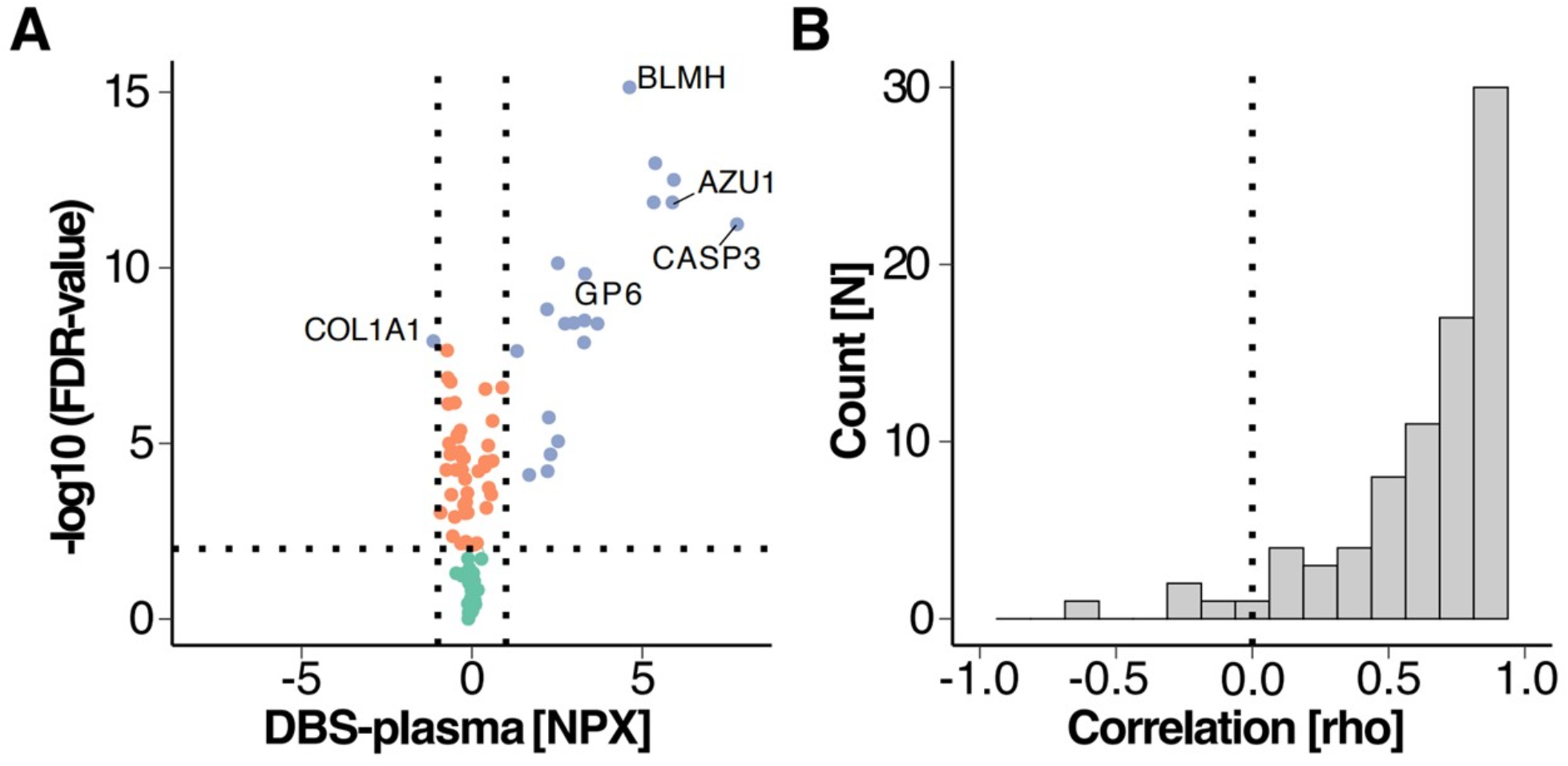
Comparison of DBS and EDTA plasma. **(A)** Volcano plot displaying difference in protein abundance between DBS and EDTA plasma for 92 proteins. The blue dots represent the proteins with FDR < 0.01 (vertical dotted line) and difference between NPX values between DBS and plasma of -1 < ΔNPX < 1 (vertical dotted lines). **(B)** Frequency of Spearman correlation coefficients for the 92 proteins. The vertical dotted line indicates rho = 0.

In summary, we observed a supportive agreement between protein profiles of DBS and plasma samples, but also noted differences in abundance levels. Discordance between the two sample types was primarily found for proteins with higher levels in DBS, which, as expected, could be due to their expression in and the presence of blood cells.

### Selection population-derived samples based on their serological immune response

During the late spring of 2020, we sent out 2000 home sampling kits to the Stockholm population to measure antibodies against SARS-CoV-2 in dried blood. Status of seropositivity was determined in 878 samples via multiple antigens (16). Using this serostatus, we selected a representative subset (*N* = 144) to perform protein profiling by PEA. We aimed at identifying components of the circulating proteome that were associated with a SARS-CoV-2 infection. The analysis was divided into two studies and driven by the objective to identify proteins associated with either no, early, current, or recent COVID-19 infections by comparing individuals showing congruent self-reporting symptoms. In study 1, we compared individuals with a strong immune response to an infection (being seropositive for both IgG and IgM as well as several SARS-CoV-2 antigens) with seronegative individuals. Both groups were matched to having have experienced similar symptoms (**Table 1**). In study 2, we investigated DBS samples to distinguish proteins associate to a recent infection - being seropositive for IgG only - with samples from donors representing the early phase of the infection - being seropositive only for IgM. Again, both groups were matched on self-reported symptoms (**Table 2**).

**Table 1.** Demographics of seropositive and seronegative population subjects (study 1).

| Serostatus | IgM<sup>+</sup>IgG<sup>+</sup><br>(seropositive) | IgM<sup>-</sup>IgG<sup>-</sup><br>(seronegative) | *P*-value* |
| --- | --- | --- | --- |
| **Participants (N)** | 41 | 37 |  |
| **Sex** |  |  |  |
| Female | 23 (56.1%) | 20 (54.1%) | 1 |
| Male | 18 (43.9%) | 17 (45.9%) |  |
| Missing | 0 (0%) | 0 (0%) |  |
| **Age group** |  |  |  |
| 20-29 | 14 (34.1%) | 13 (35.1%) | 1 |
| 30-39 | 8 (19.5%) | 7 (18.9%) |  |
| 40-49 | 6 (14.6%) | 6 (16.2%) |  |
| 50-59 | 5 (12.2%) | 4 (10.8%) |  |
| 60-69 | 7 (17.1%) | 6 (16.2%) |  |
| 70-74 | 1 (2.4%) | 1 (2.7%) |  |
| Missing | 0 (0%) | 0 (0%) |  |
| **Influenza-like symptoms** |  |  |  |
| Yes, severe | 3 (7.3%) | 0 (0%) | 0.413 |
| Yes, fever | 18 (43.9%) | 19 (51.4%) |  |
| Yes, mild | 13 (31.7%) | 13 (35.1%) |  |
| No | 4 (9.8%) | 2 (5.4%) |  |
| Missing | 3 (7.3%) | 3 (8.1%) |  |
| **Breathing symptoms** |  |  |  |
| Yes | 3 (7.3%) | 5 (13.5%) | 0.895 |
| Coughing | 5 (12.2%) | 4 (10.8%) |  |
| Hard breath | 5 (12.2%) | 4 (10.8%) |  |
| Both | 3 (7.3%) | 4 (10.8%) |  |
| No | 25 (61.0%) | 20 (54.1%) |  |
| Missing | 0 (0%) | 0 (0%) |  |

\*As confirmed by Fisher's exact test result.

**Table 2.**
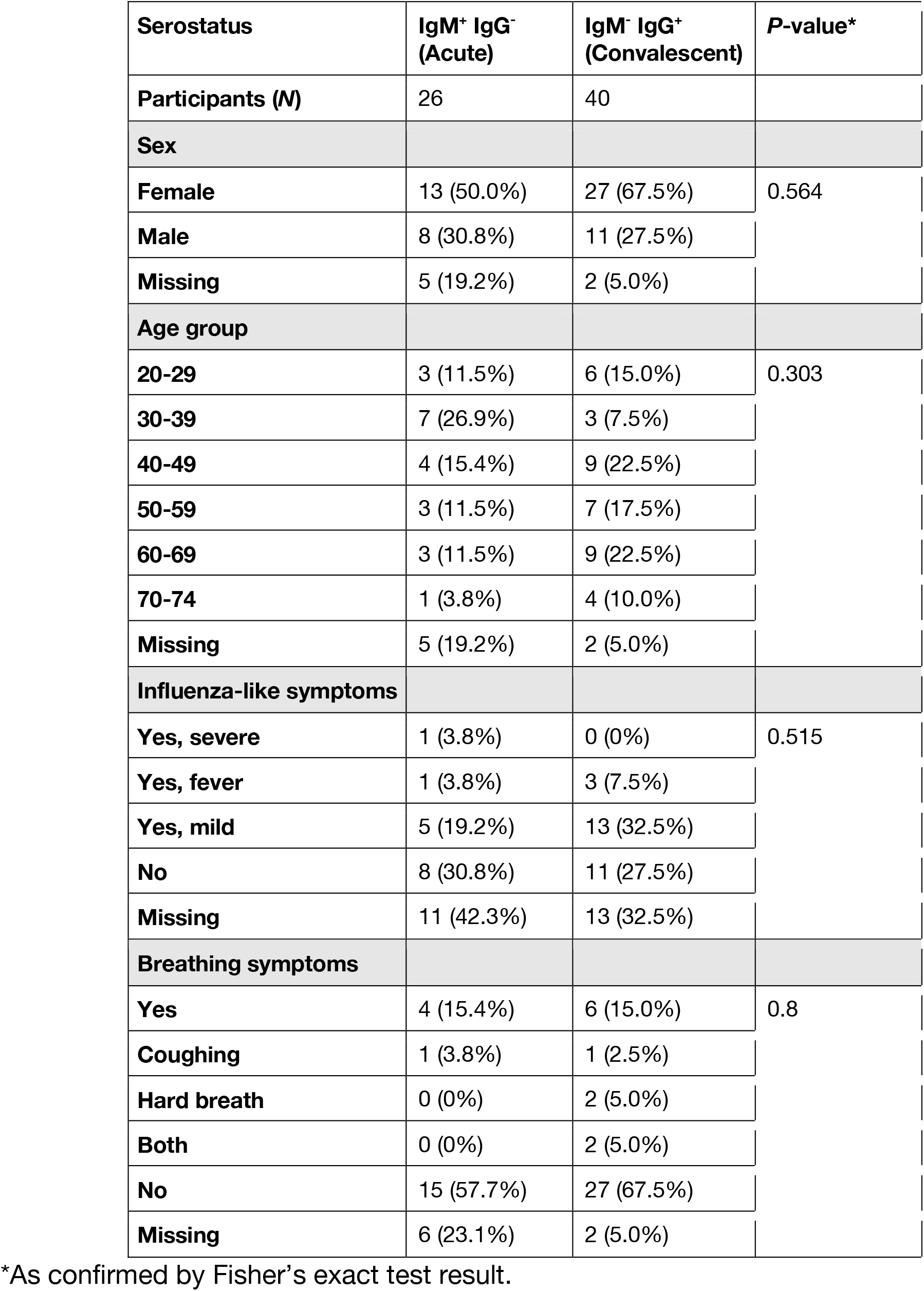
Demographics of subjects from acute and convalescent phase (study 2).

| Serostatus | IgM<sup>+</sup> IgG<sup>-</sup><br>(Acute) | IgM<sup>-</sup> IgG<sup>+</sup><br>(Convalescent) | P-value* |
| --- | --- | --- | --- |
| **Participants (N)** | 26 | 40 |  |
| **Sex** |  |  |  |
| **Female** | 13 (50.0%) | 27 (67.5%) | 0.564 |
| **Male** | 8 (30.8%) | 11 (27.5%) |  |
| **Missing** | 5 (19.2%) | 2 (5.0%) |  |
| **Age group** |  |  |  |
| **20-29** | 3 (11.5%) | 6 (15.0%) | 0.303 |
| **30-39** | 7 (26.9%) | 3 (7.5%) |  |
| **40-49** | 4 (15.4%) | 9 (22.5%) |  |
| **50-59** | 3 (11.5%) | 7 (17.5%) |  |
| **60-69** | 3 (11.5%) | 9 (22.5%) |  |
| **70-74** | 1 (3.8%) | 4 (10.0%) |  |
| **Missing** | 5 (19.2%) | 2 (5.0%) |  |
| **Influenza-like symptoms** |  |  |  |
| **Yes, severe** | 1 (3.8%) | 0 (0%) | 0.515 |
| **Yes, fever** | 1 (3.8%) | 3 (7.5%) |  |
| **Yes, mild** | 5 (19.2%) | 13 (32.5%) |  |
| **No** | 8 (30.8%) | 11 (27.5%) |  |
| **Missing** | 11 (42.3%) | 13 (32.5%) |  |
| **Breathing symptoms** |  |  |  |
| **Yes** | 4 (15.4%) | 6 (15.0%) | 0.8 |
| **Coughing** | 1 (3.8%) | 1 (2.5%) |  |
| **Hard breath** | 0 (0%) | 2 (5.0%) |  |
| **Both** | 0 (0%) | 2 (5.0%) |  |
| **No** | 15 (57.7%) | 27 (67.5%) |  |
| **Missing** | 6 (23.1%) | 2 (5.0%) |  |

\*As confirmed by Fisher's exact test result.

In study 1, 78 participants were selected based on their serological immune response against viral antigens. This included a group of 41 seropositive subjects (IgG^+^IgM^+^) and 37 seronegative individuals (IgG^-^IgM^-^) that reported a range of symptoms (**Tables 1**). There were three subjects reporting severe symptoms only in the IgG^+^IgM^+^ group, but otherwise, there were no significant differences in the self-reported symptoms between the groups. The IgG^+^IgM^+^ samples in study 1 were from participants at the peak of the immune response deemed by detachable levels of both IgG and IgM against multiple SARS-CoV-2 antigens.

In study 2, 66 participants were chosen to represent the different phases of the serological immune response against the viral infection. The stratification was based on antibodies detected against the S proteins of SARS-CoV-2. We selected 26 individuals showing signs of an early/acute immune response against the virus by being IgM seropositive only (IgG^-^IgM^+^). These were compared with 40 convalescent individuals without detectable IgM levels and being seropositive only due to IgG (IgG^+^IgM^-^) (**Table 2**). The IgG^+^IgM^-^ group was slightly older, but otherwise there were also no significant differences between the demographics and reported symptoms.

### Global analysis of proteomic profiles of population-derived DBS samples

We profiled 276 proteins associated with cardiovascular and metabolic processes such as angiogenesis, blood vessel morphogenesis, inflammation, and cell adhesion in two separated sets of samples. To begin with, we investigated the general properties and characteristics of the proteomics data without considering the serostatus categories. Our analysis of the DBS eluates revealed that a total of 260 proteins (94.2%) could be detected in > 90% of the samples. Out of 276 proteins, we found 265 that were above the limit of detection for at least 50% of the samples, and we included these in the downstream analysis. Global analyses of the protein signatures revealed no systematic differences or bias between the seropositive groups investigated in study 1 and study 2 (**Fig. S2**).

Thereafter, we evaluated the general differences in protein levels between the subjects to describe their inter-individual variability. The proteins were ranked in terms of inter quartile range (*IQR*) of the reported levels. There was a noticeable agreement in *IQR* values between the two sets (*R*^*2*^ = 0.80), hence only proteins commonly ranked at the top or bottom will be described. As illustrated in **Fig. S3** and **Data S2**, the most disperse (*IQR* > 1.25) levels were found for primarily secreted and liver proteins IGFBP1, EPCAM, and MBL2, the intestinally secreted MEP1B, as well as the intracellular liver protein NQO2. Among the least variable proteins (*IQR* < 0.15) were the intracellular BLMH, THOP1, SOD1 and BAG6 that originate from organs such as the liver or muscle tissue. Interestingly, MBL2, as protein involved in complement activation, has been studied in the context of COVID-19 (*17, 18*). BLMH, on the other hand, is highly expressed by skin tissue (*19*) and one of the proteins most differentially abundant when comparing DBS with plasma (see above).

Next, we determined the associations between protein levels and demographic data using a linear model and FDR *P* < 0.05 as cut-off (**Data S3)**. In study 1, which was built with seropositive and seronegative donors, several proteins were associated with gender (eg MMP3, SSC4D or IGFBP6) and the presence or absence of symptoms (PON3 and SELL). In study 2, where we included donors from acute and convalescent phases of the immune response against SARS-CoV-2, a smaller number of significant associations were found and only related to gender, which, reassuringly, included the shortlisted candidates MMP3, SSC4D and IGFBP6. These associations to gender are supported by RNA expression studies in tissues (*19*). The three secreted proteins originate, besides other organs, mainly from the human reproductive systems, salivary glands, and endocrine tissues. MMP3, a protease involved in collagen degeneration, has been associated with coronary heart disease and acute respiratory distress syndrome (31), and was studied in the context of COVID-19 (32).

### Circulating proteins associate with phases of the immune response against SARS-CoV-2 infections

A variety of statistical methods were used to investigate differential protein levels and correlations in relation to the SARS-CoV-2 serostatus. At first, we evaluated the heterogeneity of variance within each of the group using a Levene’s test. No significant differences (FDR *P* > 0.05) in variance were found between the groups in study 1 and study 2, however two proteins were still worth noticing. Between the groups in study 1, variance was found for the levels of the metalloprotease THOP1 (*P* = 0.0013), which had also been listed among the most proteins with the lowest IQR (**Data S2**) For the groups in study 2, the largest difference in variance was observed for the T-cell-derived cytokine CCL5 (*P* = 0.0006), which is known to be involved in the immune response. In DBS-derived data from subjects representing different serostatus and groups, there was no substantial difference in the variance of protein level.

Applied regressions analysis using LASSO were used to identify proteins related to the serostatus. The groups in study 1 and 2 were compared separately. A set of eight proteins were selected for study 1, but the model did not reveal any features for study 2. As shown in **Fig. 3**, 7 of 8 proteins had higher levels in the seropositive group, while only the primarily neutrophil-derived IgG receptor FCGR2A had a reduced abundance in relation to COVID-19. FCGR2A is known to trigger cellular response against pathogens and is involved in phagocytosis. Lower levels of FCGR2A could therefore indicate an increased SARS-CoV-2 induced clearing of immune cells.

**Fig. 3.**
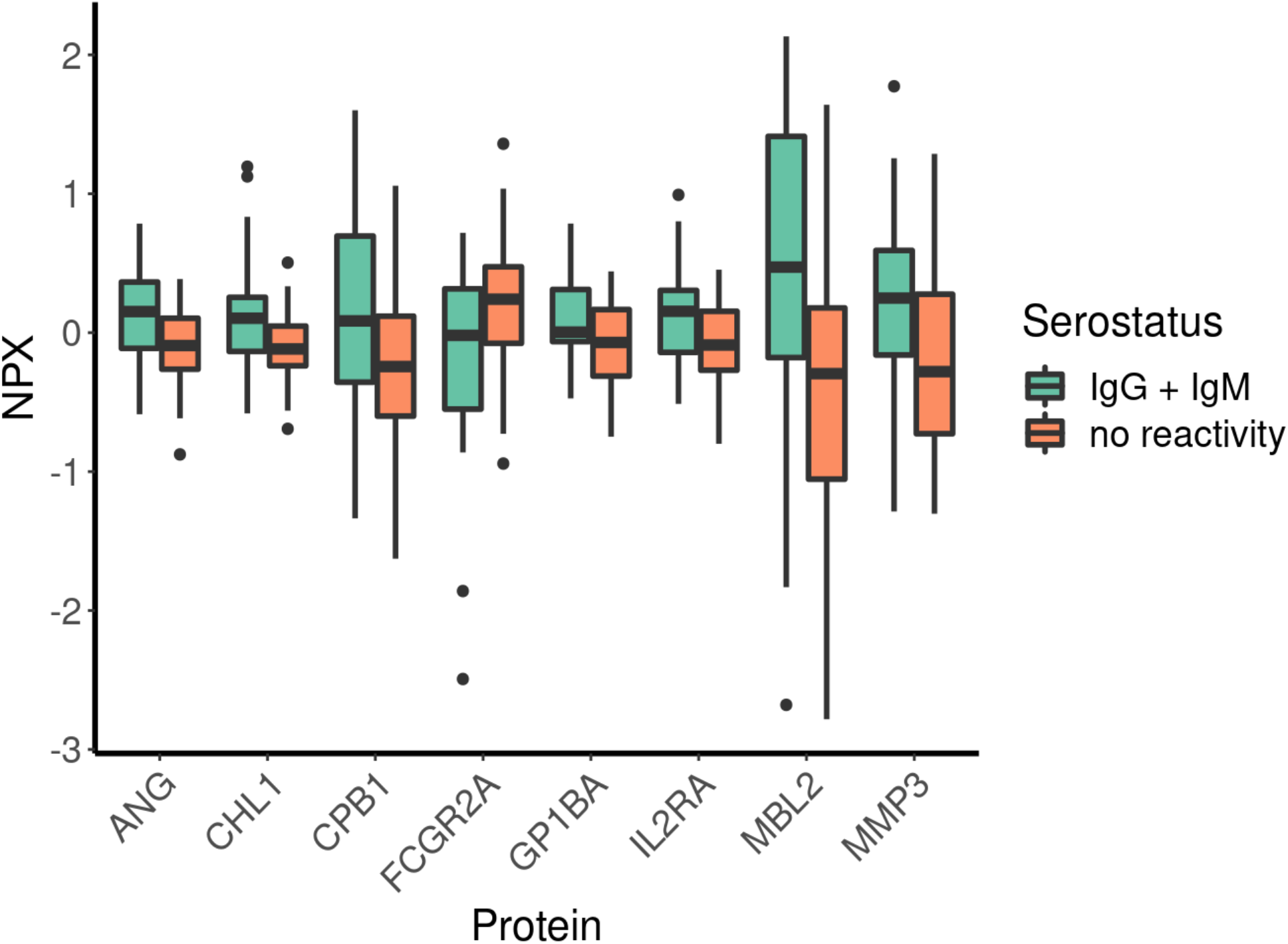
Proteins related to SARS-CoV-2 infection. LASSO analysis was applied to shortlist proteins from the comparison of seropositive and seronegative subjects in the population samples of study 1. The y-axis represents the centred and scaled data provided as NPX values.

In addition, significant differences were observed for the previously introduced MBL2 and MMP3, as well as proteins related to different physiological mechanisms. This included proteins secreted by the liver protein in stress response and angiogenesis (ANG), a brain- and B-cell-derived neurogenic protein (CHL1), a protease secreted by the pancreas (CPB1), a platelet-derived glycoprotein involved in coagulation (GP1BA) as well as a cytokine receptor related to T-cell immunity (IL2RA). Reassuringly, and as discussed later, many of these processes have been described by other in the recent literature (*20, 21*). Our observation speaks to the broad and wider-reaching physiological effects of a SARS-CoV-2 infection.

Subsequently, we compared the protein level in groups of study 1 and study 2 in relation to serostatus using a classical Kruskal-Wallis test. At a stringent cut-off of FDR *P* < 0.05, there were, however, no proteins that reached significant difference in this dichotomous comparison (**Data S4)**. The top three proteins (*P* < 0.005) in study 1 were the previously described MBL2 and MMP3, as well as VWF, well-known for its role in the coagulation system and secreted by endothelial cells. In study 2, which included subjects deemed seropositive for either only IgM (acute phase of the infection) or only IgG (convalescent phase of the infection), we observed three proteins associated with higher levels in the IgM^+^ samples (*P* < 0.025). These were PILRB, an immunoglobulin-like receptor found on granulocytes; the protein GNLY, which is secreted by NK- and T-cells and involved in anti-microbial processes; as well as LILRB1, which is another immunoglobulin-like receptor found on monocytes. Univariate statistics provided insights about the effects of a SARS-CoV-2 infection on the complement systems. This pointed at proteins from the immune system to differentiate the physiological effects occurring during the acute and convalescent phase of the immune response.

To compare proteins levels more directly with the actual levels of IgG and IgM previously reported for the S, RBD and N proteins of the virus (*16*), we conducted correlation analysis. Even though none of the relationships reached statistical significance (FDR *P* < 0.05), we observed milder but noteworthy correlations (*rho* ± 0.35; *P* < 0.001). In study 1, there were negative correlations between FCGR2A, and IgG levels detected for RBD (*rho* = -0.38; *P* = 0.0006) and S (*rho* = -0.32; *P* = 0.004). MMP3 levels were found to correlate with IgG levels for S (*rho* = 0.37; *P* = 0.0008) and RBD (*rho* = 0.35; *P* = 0.002), and similar trends and relationships were determined for MBL2 and VWF. For IgM in study 1, we observed circulating CHL1 to correlate with levels reported for RDB (*rho* = 0.44; *P* = 0.00007) and S (*rho* = 0.38; *P* = 0.0006), and similarly for previously mentioned MBL2, VWF, GP1BA, ANG. Intriguingly, a mild negative correlation was observed also between FCGR2A, and IgM levels detected for S (*rho* = -0.26; *P* = 0.02) and RBD (*rho* = -0.23; *P* = 0.04) In study 2, none of the correlations reached the nominal p-value threshold **(Data S5)**. We explained the lack of significant correlations by the lower resolution of the serology analysis, imbalanced numbers of samples per group, and for the IgG^+^ group the unknown time difference between infection and sampling.

### Changes in protein-protein correlation networks occur between the phases on the immune response

We demonstrated the capability that DBS proteome profiling can reveal valuable insights about protein levels associated with different serological stages of COVID-19. To learn more about mechanistic alterations induced by a SARS-CoV-2 infection, we conducted correlation analyses of protein levels within each of the four serological groups. As depicted in heat maps in **Figs. 4A** and **B**, different relationships between the protein correlations could be observed.

**Fig. 4.**
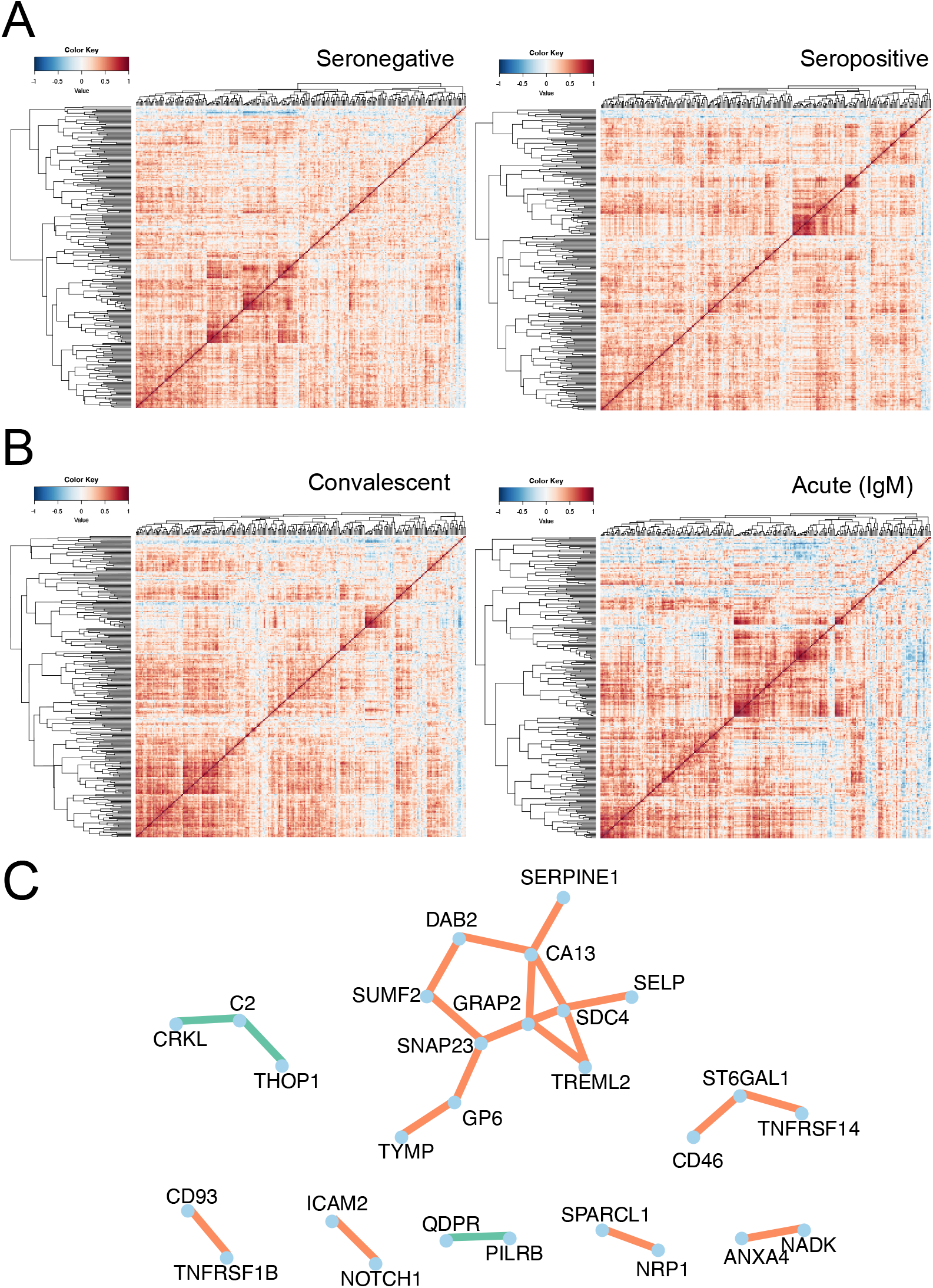
Protein correlation landscape in serostatus groups. **(A-B)** The heat maps reveal the inter-protein correlations observed within the four serological groups from study 1 and 2. **(C)** A representation of the strongest protein-protein correlations differing in individuals from the acute and convalescent phase of the infection (study 2)

To identify which of the observed correlations differed between the serostatus groups of study 1 and study 2, we performed differentiation co-expression analyses. Each protein pair was first categorized based on the direction of correlation, marked by (0), (+) or (-), see **Data S6**. Among the nearly 35.000 tested pairs, > 55% were classified to be below a default correlation threshold (nominal *P* < 0.05) within each serostatus group, marked as (0/0). Among the remaining pairs, > 97% were positively correlated in both groups (+/+), while only ∼1.3% were negatively correlated (-/-). We further limited the search space to differential protein-protein correlations with *P* < 0.01.

For study 1, comparing seropositive with seronegative subjects, two protein pairs with changes in their correlation trends (+/+) emerged: COMP-CCDC80, a pair of extracellular matrix proteins found fibroblast and involved in cell adhesion, as well as GRN-CXCL16, which are two cytokines secreted by monocytes. In study 2 (IgM^+^IgG^-^/ IgM^-^IgG^+^), three protein pairs with opposite trends (+/-) and 20 pairs with different steepness of the trends (+/+) emerged. Most noticeable for (+/-) were differences in correlations for the secreted complement protein C2 with either the metalloprotease THOP1 or the intracellular adaptor protein CRKL. Among the remaining 20 (+/+) pairs, all reported an elevated inter-protein correlation within the IgM^+^IgG^-^ group, which represents subjects from the early phase of the infection. In these 20 pairs, we found an overrepresentation and network containing the secreted proteoglycan SDC4, the vesicle transporter SNAP23 from granulocytes, the intestinal carbonic anhydrase CA13, and GRAP2, an adaptor protein from lymphoid tissues (**Fig. 4C**).

Noteworthy here was SDC4, a protein originating from liver, lung, and kidney and that, like ACE2, has recently been linked to the cellular uptake of SARS-CoV-2 virus (*22*). Previous studies have revealed anti-inflammatory functions in patients with acute pneumonia (*23*).

In summary, differential correlation analysis revealed additional insights about coordinated molecular changes occurring in COVID-19 subjects, who had been stratified by serostatus. Processes related to the immune system, cell adhesion but also cellular virus entry could be identified in DBS samples collected from a general population.

## Discussion

The COVID-19 pandemic highlighted the need for fast and effective strategies for population health surveillance, in respect to infectious diseases spreading and the short- and long-term consequences on a heterogeneous population (*7*). To fully unlock the possibilities offered by advanced methodologies, new and precise analytical capabilities linked to patient-centric sampling at home, such as dried blood spots (DBS), will become an essential cornerstone (*24*). We applied volume-controlled DBS sampling when the first wave of the pandemic hit Sweden and collected > 800 blood samples from random households. Based on serological analyses, we chose to investigate > 250 circulating proteins in DBS samples for their associations to groups of participants representing different phases of the SARS-COV-2 infection. To our knowledge, this is the first proteomics survey conducted in a general population. The integrity of the sample type enables the use of home sampling to generate molecular data of high quality and with large cohorts.

Proteomics has shown to deliver important insights about COVID-19 in clinical plasma samples (*25*). We chose PEA for the proteomics analyses due to their low sample volume requirement and multiplexed design paired with high sensitivity and specificity. PEAs have previously been successfully applied for the quantification of proteins collected in filter paper-based DBS samples obtained in clinical or professional settings. There, the technology showed convincing capabilities to detect proteins even decades after sampling (*26*) and good correlation with the measurements obtained from samples collected by venous blood draw (*27, 28*). However, it is well known that intra- and inter-individual variability in hematocrit levels or other blood cell counts may affect the dispersion of whole blood on paper-based DBS collection matrices. The use of a volumetric device not only guarantees a higher level of robustness of analysis but also allows the automation of the procedure of extraction, avoiding manual punching and increasing the throughput of analysis. Our data confirmed robustness and reproducibility on protein quantification (*CV* < 10%) and a convincing concordance between protein profiles in EDTA plasma collected by venous blood draw and DBS collected by finger pricking (*rho* > 0.7). This also confirmed that chosen dilutions of the samples provided an equivalent number of proteins as a liquid sample.

As predicted, proteins with a higher abundance in DBS samples originated from skin, intracellular, and blood cells (e.g. GP6, AZU1, CASP3), suggesting cell lysis as the main reason. Variability analysis revealed cellular proteins such as BLMH, THOP1, SOD1 and BAG6 are among the least variable proteins (*IQR* < 0.15). This indicates comparable counts of blood cells were trapped in the discs. We consider the presence of blood cells in DBS sample an intrinsic advantage for expanded global blood profiling.

Circulating proteins serve as important sentinels for specific pathway activation and organ status, not only in monitoring disease progression and response to therapy in severe hospitalized cases, but eventually also for the investigation of the causes behind long term symptoms experienced by mild COVID-19 patient (*4*). Despite most patients with COVID-19 overcome the disease experiencing only mild to moderate symptoms, severe patients may develop pneumonia, acute respiratory distress syndrome (ARDS) and in general a multi-pathologic and complex clinical picture. The most known pathologic features include cytokine release syndrome or “cytokine storm”, lung, cardiovascular and kidney dysfunction, increased thrombotic risk, and down-regulation of adaptive cellular immunity (*29*). Altered levels of multiple circulating proteins have been observed particularly in severe patients, including the inflammatory IL-6 and CRP being elevated in severe and hospitalized patients (*30*). Overproduction of pro-inflammatory cytokines causes a dysfunctional balance between the mechanisms of pro-coagulation and anti-coagulation which will result in disseminated micro-thrombosis and cause multi-organ failure, and in severe cases, death (*2, 3*). Furthermore, high levels of cardiac biomarkers such as Troponin, BNP and MBL2 were identified as strong predictor of mortality (*31, 32*).

The comparison between IgG^+^IgM^+^ and seronegative individuals highlighted some interesting aspects related to the feature of mild/asymptomatic COVID-19 pathology. MBL2, a protein associated with ICU mortality (*22*) was here found at high levels also in individuals who had a mild/asymptomatic disease when compared to healthy seronegative individuals. No difference was observed between subjects representing the acute (IgG^-^IgM^+^) and convalescent (IgG^+^IgM^-^) phases of the immune response, suggesting that the protein may circulate at elevated levels also after recovery. A similar trend as for MBL2 was observed for MMP3 and PRSS, two proteins involved in antimicrobial humoral response and VWF an important factor in blood coagulation. In the group of seropositive individuals, we also identified a blood protein signature associated with cell-mediated immune response and tissue damage, mechanisms lying behind severe COVID-19 (*20, 33*)

We also performed a small pilot study using longitudinal DBS samples (**Supplementary Text**) to generate plasma protein profile of a COVID-19 patient experiencing moderate symptoms (fever, anosmia, ageusia and dry cough). The immunoglobulin profiles suggest that the individual had reached the peak of acute phase at day 15, starting the convalescence phase (IgG^+^IgM^+^, with constant IgG and decreasing IgM levels) (**Fig. S5C**) (*16, 34*). Markers of severe and acute COVID-19, such as IL2RA (*35*), which were found elevated in seropositive individuals in study 1 (**Fig. 3**), showed a decreasing trend over time in the convalescent donor. IL2RA, together with IL18 and IL18BP, correlated in COVID-19 patients with troponin levels and is therefore considered a marker of cardiovascular damage involvement (*36*). Proteins such as CRTAC and CHRDL2, suggested to play a role in osteoblast differentiation and maturation (*37*), showed instead an inverse trend. The observed increase in levels of circulating CHRDL2 may support recent findings of an active role of bone marrow as immune regulatory organ and indicate active proliferation of cells involved in adaptive immune response (*38*)

An inherent limitation of studying DBS samples is the need for very sensitive methods for quantification. (*9*). In this study we chose to focus on stable proteins occurring at medium to high abundance levels in the circulation. Indeed, the recommended dilution for plasma for the three assay panels used is 1:100, 1:20, or 1:2025. Such dilutions could be easily coupled to the blood dilution implicit in the procedure of protein elution from DBS (see Material and Methods). Consequently, next efforts aim to establish procedures to quantify proteins of lower abundance, such as IL6 or TNF, as well as other inflammatory biomarkers discussed in the literature. Moreover, since the chosen method build on pre-selected panels of proteins, we could have missed some relevant markers described covered in current COVID-19 literature (*39*).

As population samples were collected in an anonymous manner from a set of random households, no follow-up of the participating donors will be possible and new studies must be designed where additional samples and clinical information can be collected over a longer period. Even though we observed associations between serostatus and proteins, there could be unknown factors, such as BMI, medication, travel, or socio-economic factors as well as lifestyle contributing to the difference of this analysis. Such factors will be necessary to take into consideration in future studies to further evaluate the results presented here.

The presented study focused on understanding circulating proteins of non-hospitalized individuals during the first wave when no vaccines or extensive testing capabilities were available. Blood samples were collected from random persons and none of these reported being diagnosed through PCR tests. According to the questionnaire data (**Tables 1**. and **2**.), the majority were asymptomatic or only reported milder symptoms. Uncertainty remains about the time-point of infection with respect to the point of sampling, adding further heterogeneity to the population. With additional knowledge of clinical information, future studies will therefore have the possibilities to understand the immune response in SARS-CoV-2 patients in more detail, and to enrol patients from or after intensive care and use home sampling to follow these over a longer period. Focussing on recovery trajectories of IgG^+^IgM^-^ groups could provide valuable information on their health states, and identify proteins related to recovery and long COVID-19.

Proteomics analyses of circulating proteins in population-based DBS samples collected during the first wave of the COVID-19 pandemic provided valuable insights into the molecular effects of a SARS-CoV-2 infection. The quality of the data and the concordance of our findings with clinical studies and a growing literature supports the utility and potential of our approach as a viable option for assessing states of public health.

## Materials and Methods

### Samples and sampling

#### Comparison of DBS and EDTA plasma

Venous and capillary blood samples were collected from volunteers (*N* =12) among personnel at a healthcare center in the Stockholm region as previously described (16). In short, venous blood was collected through venipuncture into EDTA blood collection tubes (K2E K2EDTA Vacuette tube, #454410, Lot# A19104MX, Greiner Bio-One). The tube was centrifuged, and the blood plasma was collected and stored at –20 °C until further use. Capillary blood samples were obtained by finger-pricking and applying blood droplets onto a quantitative DBS sampling card (qDBS, Capitainer AB, Stockholm, Sweden) according to the supplier’s instructions. The qDBS cards were stored at room temperature until heat treatment prior to extracting the blood-filled discs.

#### Population study

Capillary blood samples from the general population were obtained by cold-mailing home-sampling kits (MM20-009-01, Capitainer AB, Sweden) to 2000 randomly selected individuals (20-74 years old) in metropolitan Stockholm (**Tables 1**. and **2**.) during April 2020 together with a questionnaire, as described previously (16). Individuals who volunteered to participate in the study were asked to perform self-sampling according to the instructions and return the filled sampling card, questionnaire, and consent form by regular mail. All cards were barcoded and stored at room temperature until use, or as stated otherwise. All anonymous blood donors gave informed documented consent. The study was approved by the regional ethical board (EPN Stockholm, Dnr 2015/867-31/1) and the Swedish Ethical Review Authority (EPM, Dnr 2020-01500). The data on serostatus of the collected samples was determined via multiple antigens and described previously (16).

#### Longitudinal study

DBS samples were collected from an anonymous volunteer during the early recovery phase of a PCR-confirmed SARS-CoV-2 infection. The samples were collected in five separate occasions, during week 2 to 5 after symptom onset.

### Affinity proteomics assays

#### DBS eluates preparation

The eluates were prepared as previously described. In short, the blood sampling cards were inactivated in an oven (UN55m, Memmert GmbH) at 56°C for 60 min prior to ejecting the discs into separate wells of a flat bottom 96-well plate (#734-2327, VWR). The content of the discs was eluted using 100 μl of PBS with 0.05% Tween20 (#97062-332, VWR) and protease inhibitor cocktail (#04693116001, Roche) followed by gentle shaking (170 rpm) for one hour at room temperature. The plates were then centrifuged for 3 min at 3000 rpm (Allegra X-12R, Beckman Coulter Inc.) and 70 μl supernatant was transferred into a PCR plate (#732-4828, VWR). Sample eluates were stored at -20°C until analysis.

#### Proximity extension assays

Multiplexed protein analysis was performed at SciLifeLab’s Unit of Affinity Proteomics-Stockholm using Olink panels Cardiovascular III (Product No 95611, Lot No B01116), Metabolism (Product # 95340, Lot # B01109), and Cardiometabolic (Product No 95360, Lot No B02504) according to manufacturer’s instructions (Olink Proteomics AB). In brief, EDTA plasma samples were diluted according to the manufacturer’s instructions to, depending on the panel, 1:100, 1:20, or 1:2025. We estimated a dilution of plasma in DBS eluates of 1:20. Eluates were then diluted 1:5, 1:1, or 1:101, respectively. For each panel, samples were incubated with 92 pairs of oligonucleotide-labelled antibodies simultaneously. Upon target recognition, the oligonucleotides in the antibody pairs are brought in close proximity allowing for hybridization and DNA polymerization. Reporter sequences were quantified using a microfluidic real-time PCR instrument (Biomark HD, Fluidigm), data were processed using the software NPX Manager (v.2.1.0.224 and v2.2.0.288, Olink Proteomics AB) and protein relative quantification was reported as normalized protein expression (NPX) values.

### Statistical analyses

Data analysis and visualizations were performed with the statistical software R version 3.6.0 (*40*).

#### Comparison of plasma and DBS

Paired t-tests were performed using the “t.test” function from the “stats” R package (v3.6.0), and the “p.adjust” function from the same R package were used to calculate the FDR values.

#### Global analyses

In total, 265/276 proteins were delivered data above the limit of detection for > 50% of all samples included in the analysis. Three outlier samples that were identified by PCA and hierarchical clustering (Euclidean distance) as having consistently lower NPX across all three panels. As this is likely due to technical differences during the sample processing, the three DBS samples were excluded from the analysis.

### Data normalization

NPX data from two experiments (study 1 and study 2) were bridge-normalized and combined into one data set. Bridge normalization was performed by:

1. Calculating the difference between each protein per paired bridge sample
2. For each protein, taking the median of this differences across the bridge samples
3. Adjusting each protein with the median in one of the experimental batches.

#### Lasso regression

The analysis was performed using the R package “glmnet” (version 4.1) (*41*) to find features that were informative for serostatus in each data set. The regression was performed 10 times for each data set and the intersection of the chosen features across the 10 regressions was chosen as the informative features.

#### Association tests

Linear regression models adjusted for age-group and sex were applied per protein profile. Residuals obtained from the linear models were then used in the analysis. Kruskal Wallis rank sum test was performed for association tests between protein data and sample information, including age group (“Age_grp”), sex, seropositivity (“Analysis_group”), symptoms (“Symptom_simplified”), breath (“Breath_simplified”), population batch and assay plate (“Sample_plate”) (Supplementary data file 4). Here, breath and symptom questionnaire information were converted into binary data (yes/no). All *P*-values were FDR adjusted for multiple correction using the Benjamini-Hochberg method.

#### Correlation analysis

Spearman’s *rho* statistic was used for estimating correlations, unless otherwise specified. Correlations between the determine protein levels and IgG or IgM antibody levels detected against S, RBD, and N antigens in population samples (separately per set, *N* = 78 and *N* = 66) were computed using the “corr.test” function of the “psych” R package (version 1.9.12.31). Two-tailed P-values were adjusted for multiple correction using the Benjamini-Hochberg method.

#### Differential correlation analysis

Differential correlation analysis was performed, using the “ddcorAll” function of the “DGCA” package (version 1.0.2) (*42*), to compare protein-protein correlation between different serostatuses within each study. Two-sided p-values for the correlation differences were generated from Z-scores of the Spearman correlations (*43*). A network plot showing pairs of proteins with *P*-values below 0.01 for correlation within each serostatus and the difference between statuses was generated for study 2 using the “ggnet2” package (included in the “GGally” package version 2.1.2) (*44*) Resulting correlation values were visualised as heatmaps with rows and columns clustered with hierarchical clustering using correlation distance.

#### Longitudinal analysis

Longitudinal samples were included in duplicate. Protein profiles were scaled and centered, and the mean value between replicates was calculated. The correlation between each replicated protein was assessed. The mean replicate values of all 276 proteins were used in downstream analysis. Self-Organizing Tree Algorithm (SOTA) cluster size *N* = 9 was applied on the protein data.

## Data Availability

Anonymised data will be deposited at the SciLifeLab Data Repository (https://scilifelab.figshare.com) and access to the data can be granted for validation purposes and upon reasonable request to the corresponding authors. Analysis codes will be made available upon publication at https://github.com/Schwenk-Lab.

## Acknowledgments

We first like to thank all anonymous volunteers contributing with blood samples to our study. We thank Mun-Gwan Hong for the statistical support, Vincent Albrecht, and the members of the Nilsson Lab for fruitful discussions. We thank Olink Proteomics AB for their support. Figure 1 was created using BioRender.com.

## Author contributions

A.B., M.D., and C.M. performed experiments. T.D.C., L.D., A.B., and C.E.T. performed data analysis. O.B., N.R., J.M.S. collected clinical specimens and data and curated clinical data. J.M.S., N.R. and C.F. conceptualized the study and supervised the staff. C.F., T.D.C. and J.M.S wrote the original draft of the manuscript, and all authors reviewed and edited the manuscript.

## Data and materials availability

Analysis codes will be made available upon publication at https://github.com/Schwenk-Lab. Data availability: Anonymized data will be deposited at SciLifeLab Data Repository (https://scilifelab.figshare.com) and access to the data can be granted for validation purposes and upon reasonable request to the corresponding authors.

